# Using a novel echo-marker to identify high mortality risk patients with moderate aortic stenosis

**DOI:** 10.1101/2021.10.21.21265320

**Authors:** Chieh-Ju Chao, Pradyumma Agasthi, Timothy Barry, Marlene Girardo, Amith R. Seri, Anusha Shanbhag, Yuxiang Wang, Steven J. Lester, Said Alsidawi, William K Freeman, Tasneem Z. Naqvi, Mackram Eleid, David Fortuin, John P. Sweeney, Peter Pollak, Abdallah El Sabbagh, Kristen Sell-Dottin, David Majdalany, Carolyn Larsen, David R Holmes, Jae K. Oh, Christopher P. Appleton, Reza Arsanjani

## Abstract

**Background:** Recent studies have shown that patients with moderate aortic stenosis and reduced left ventricular ejection fraction may benefit from earlier intervention instead of periodic surveillance. Identifying patients at higher risk is therefore warranted concerning the possibility of expanding aortic valve replacement indication.

**Objective:** We aim to investigate the usefulness of a novel echo-marker, augmented mean arterial pressure (AugMAP), in identifying high-risk patients with moderate aortic stenosis.

**Methods:** Adult patients with moderate aortic stenosis (aortic valve area 1.0-1.5 cm^2^) at Mayo Clinic sites in 1/2010-12/2020 were identified. Baseline demographics, echocardiography, and all-cause mortality data were retrieved. Patients were grouped into higher and lower AugMAP groups using a cutoff of 80 mmHg for analysis. Kaplan-Meier and Cox regression analyses were used to assess the performance of AugMAP.

**Results:** A total of 4,563 patients with moderate aortic stenosis were included. The mean age was 73.7±12.5 years and 60.5 % were male. The median follow-up was 2.5 years, and 36.0% of patients died. The mean LVEF was 60.1 ± 11.4%, and the mean AugMAP was 99.1 ± 13.1 mmHg. Patients in the lower AugMAP group, with either preserved or reduced LVEF, had significantly worse survival performance (all p< 0.0001). Multivariate Cox regression showed that AugMAP was independently associated with all-cause mortality after adjusting for age, sex, and LVEF (HR: 0.99 per unit increase, 95%CI: 0.978-0.996, p=0.01).

**Conclusion:** AugMAP is a simple and effective echo-maker beyond LVEF to identify high-risk moderate aortic stenosis patients who may benefit more from earlier intervention.

**Condensed abstract:** Patients with moderate aortic stenosis and reduced left ventricular ejection fraction may benefit from earlier intervention. We aim to validate the usefulness of a novel echo-marker, augmented mean arterial pressure (AugMAP), in identifying high-risk patients with moderate aortic stenosis. AugMAP can identify patients at higher mortality risk within the first two years after the diagnosis of moderate aortic stenosis, regardless of LVEF. AugMAP was also independently associated with all-cause mortality after adjusting for age, sex, and LVEF. AugMAP is a simple and effective novel echo-maker beyond LVEF to identify high-risk moderate aortic stenosis patients who may benefit from earlier intervention.

## Introduction

Aortic stenosis (AS) is one of the most common valvular abnormalities in developed countries affecting approximately 5% of individuals older than 65 years old (1). The current ACC/AHA guidelines recommend surgical or transcatheter aortic valve replacement for appropriate patients with severe symptomatic AS(1). In contrast to the explicit suggestions for severe AS, moderate AS is a relatively gray zone lacking clear evidence to guide the practice. Overall, before reaching the stage of severe symptomatic aortic stenosis, the current guidelines suggest periodic monitoring with transthoracic echocardiography unless other concomitant cardiac surgery is indicated(1). However, there is growing evidence that patients with moderate aortic stenosis may have a worse longer-term survival. In addition, it appears that patients with moderate aortic stenosis and reduced LV ejection fraction may benefit from earlier intervention(2–4). Jean et al. demonstrated that transcatheter aortic valve replacement (TAVR) led to better clinical outcomes in patients with moderate aortic stenosis and heart failure with reduced ejection fraction(4). Furthermore, the ongoing TAVR-UNLOAD trial is investigating the efficacy of TAVR intervention in this patient population(5).

Foreseeing the possibility of an expanded indication for aortic valve placement in patients with moderate AS, it is essential to determine the optimal timing of intervention and predict post-procedure outcomes in this population. Our group previously identified a new parameter, augmented mean arterial pressure (AugMAP), which supersedes the performance in predicting 1-year mortality after TAVR(6). AugMAP is derived from noninvasive blood pressure measurements and mean aortic valve gradient determined by continuous-wave Doppler, as a surrogate marker of cardiac contractile reserve in patients with aortic stenosis. We hypothesized that augmented mean arterial pressure could predict the primary endpoint of all-cause mortality in patients with moderate aortic stenosis. We anticipate this new parameter has the potential to guide future studies to select proper candidates for aortic valve replacement in patients with moderate aortic stenosis.

## Methods

### Study population, baseline demographics, and clinical data

All patients aged ≥18 years with a diagnosis of aortic stenosis from January 1st, 2010, to December 31st, 2020, at three Mayo academic medical centers located in Rochester, MN, Phoenix, AZ, and Jacksonville, FL were identified. Patients who had prior aortic valve replacement procedure(s) were excluded. Baseline demographics, medical history, lab data, and follow-up data (including subsequent aortic valve surgical/ transcatheter intervention and all-cause mortality) were extracted from the electronic medical record. The first qualifying echocardiography during the time of the study was considered as the beginning of the study.

Moderate aortic stenosis was defined as aortic valve area in the range of 1.0-1.5 cm^2^ determined by continuous-wave Doppler echocardiography using continuous velocity equation. Patients with missing values in augmented mean arterial pressure or follow-up data were excluded to ensure the consistency of survival assessment. The Institutional Review Board at Mayo Clinic approved the study protocol, and all the patients provided research authorization to utilize their medical information.

### Transthoracic echocardiography

Transthoracic echocardiography with 2-dimensional imaging and Doppler were performed pre-procedure using commercially available ultra-sound scanners (Philips iE33; Phillips Medical Systems, Andover, MA, USA; GE Vivid E9, GE Healthcare, Milwaukee, WI, USA). All echocardiograms were interpreted according to the current society guidelines(7–11). Offline measurements of the images were obtained using ProSolv Cardiovascular Analyzer 3.0 (ProSolv Cardiovascular Inc., Indianapolis, IN, USA).

### Calculation of augmented mean arterial pressure and valvulo-arterial impedance

The augmented blood pressure calculation formulas are stated as below (12):

Augmented SBP1(AugSBP1): Mean aortic valve gradient (mean AVG) was added to systolic blood pressure **(Equation 1)** and augmented MAP1(AugMAP1) was calculated by replacing the SBP with augmented SBP1 in the MAP formula **(Equation 2);** Valvulo-arterial impedance (Zva) was calculated according to the standard formula by dividing the sum of the systolic blood pressure and mean transvalvular gradient by stroke volume index (SVI)(**Equation 3**)(13).

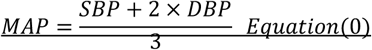

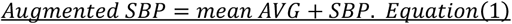

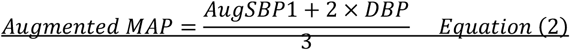

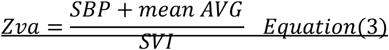

### Statistical analysis

A cutoff value analysis was performed to determine the optimal AugMAP cutoff for predicting all-cause mortality during the follow-up period. In addition, two-year all-cause mortality was used as another endpoint to investigate the ability of AugMAP in detecting patients who are at risk of early mortality. The surv_cutpoint function to the survimer R package was used to determine the optimal cutoff point for AugMAP based on the maximally selected log-rank statistics (R version 4.0.3). The patients were then divided into higher AugMAP (> cutoff) and lower AugMAP (≤ cutoff) groups and analyzed accordingly. Continuous variables were summarized as mean ± standard deviation, and the differences among groups were evaluated with the Mann-Whitney U test. Categorical variables were expressed as counts and percentages, and differences among groups were assessed with the Chi-square test. All the two-group comparisons were summarized as higher AugMAP group vs. lower AugMAP group if not otherwise specified. Kaplan-Meier curve and Cox regression were used for survival analysis. Univariate and multivariate Cox regression analyses were used to assess the association of each parameter and all-cause mortality. In multivariate Cox regression analysis, the models were adjusted for age, sex, and LVEF. A p-value of less than 0.05 was used as the cutoff of statistical significance for all the hypotheses. All of the rest analyses were performed in Python version 3.7.10.

## Results

### Patient population

A total of 4,563 patients were included in the final analysis. The mean age was 73.7±12.5 years old, 60.5 % were male, and 94.4% were white. All-cause mortality endpoint occurred in 1643 (36.0%) patients during the follow-up period (median 2.5 years, interquartile range (IQR) 0.57 years to 5.7 years). The mean LVEF was 60.1 ± 11.4%, and the mean AugMAP was 99.1 ± 13.1 mmHg (IQR, 90.3 mmHg-107.7 mmHg). There were 3,964 patients who had normal LVEF (≥ 50%) and 599 patients who had reduced LVEF (<50%). In our patient cohort, 1,274 patients underwent an aortic valve replacement procedure (1,244 surgical aortic valve replacement and 30 transcatheter aortic valve replacement) during the follow-up period; the median time from the first TTE to AVR was 2.08 years (IQR: 0.5-4.1 years).

### The optimal cutoff for AugMAP

Using all-cause mortality during the study period, the optimal cutoff for AugMAP was 87.3 mmHg (**Figure 1A**), when 2-year all-cause mortality was used, the optimal cutoff became 80.0 mmHg (**Figure 1B**). For better visualization of the survival difference, AugMAP of 80 mmHg was used as the cutoff for group and Kaplan-Meier analysis. The Kaplan-Meier survival curve plots using 87.3 mmHg as cutoff are provide in **Supplement Figure 1**. Using AugMAP of 80 mmHg as the cutoff, there were 4,256 patients in the higher AugMAP group and 307 patients in the lower AugMAP group.

**Figure 1.**
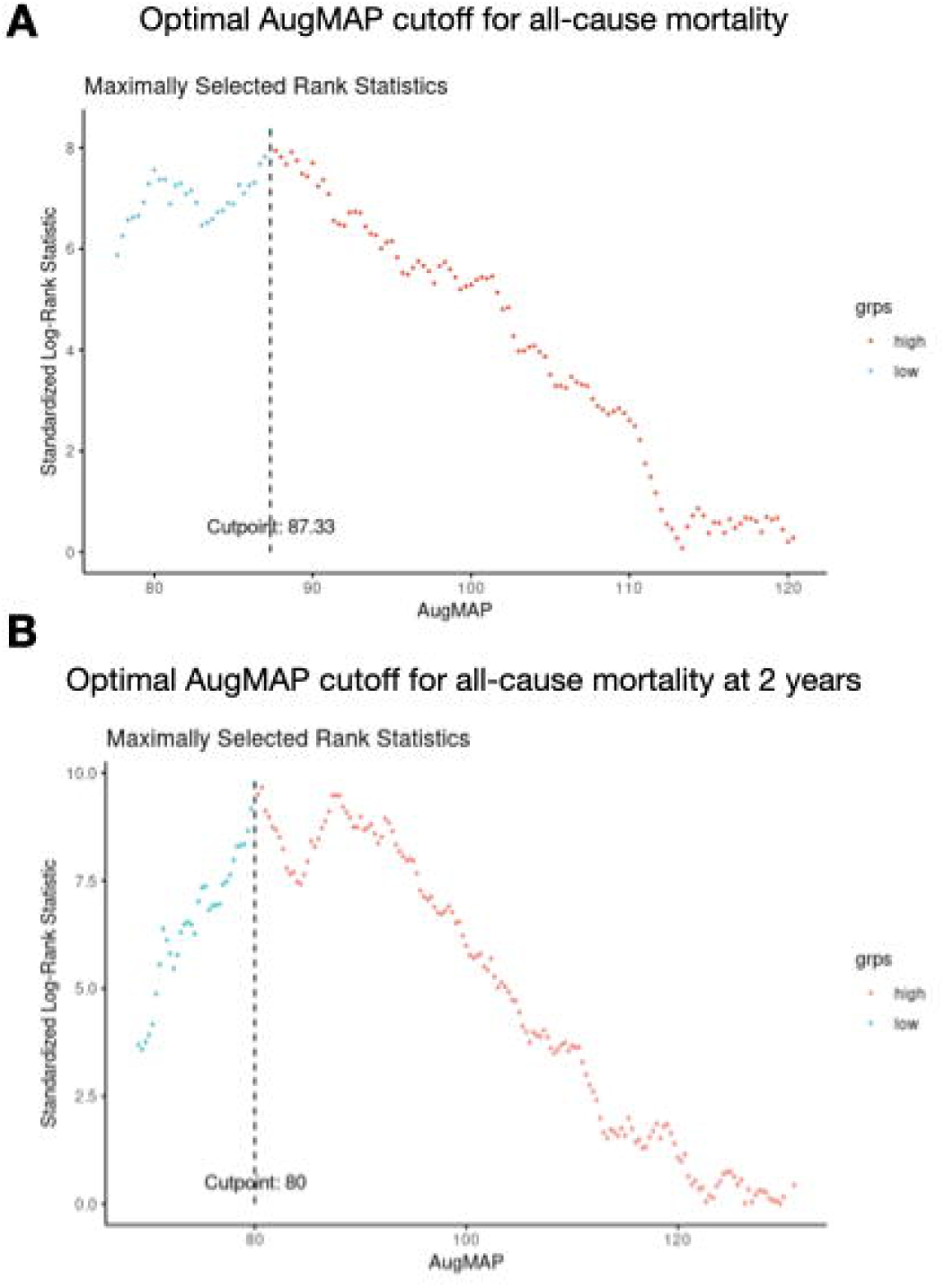
Plots of cutoff value analysis for the optimal AugMAP cutoff for predicting all-cause mortality. **Panel A**. an AugMAP of 87.3 mmHg was deemed to be the best cutoff for all-cause mortality during the follow-up period. **Panel B**. When considering 2-year mortality as the endpoint, the optimal cutoff shifted to a lower value of 80.0 mmHg. Horizontal axis: AugMAP (mmHg), vertical axis: standardized log-rank statistics.

### Baseline characteristics of the higher and lower AugMAP groups

In the lower AugMAP group, there were higher ratios of patients with a diagnosis of CAD (23.4% vs. 39.1%, p<0.0001), congestive heart failure (17.9% vs. 43.3%, p<0.0001), chronic kidney disease (20.7% vs. 39.1%, p<0.0001), use of beta-blocker (25.9% vs. 38.1%, p<0.0001) and diuretics (24.2% vs. 44.3%, p<0.0001). In contrast, we did not observe significant difference regarding the presence of hypertension (56.9% vs. 52.8%, p=0.36), diabetes (20.1% vs. 21.8%, p=0.77), nor use of calcium channel blockers (23.2% vs. 16.9%, p=0.04), ACE inhibitors (4.6% vs. 6.2%, p=0.45) and ARBs (13.2% vs. 10.7%, p=0.47).

In terms of AugMAP and other echocardiographic parameters, the higher AugMAP group had a mean AugMAP of 100.9 ± 11.5 mmHg vs. 74.1 ± 5.3 mmHg (p< 0.0001). All the components of AugMAP were significantly lower in the lower AugMAP group (SBP: 133.2 ± 19.5 mmHg vs. 98.7 ± 13.8 mmHg, DBP: 72.1 ± 10.7 vs. 51.3 ± 7.4 mmHg, mean aortic valve gradient 25.2 ± 8.3 mmHg vs. 21.0 ±7.4 mmHg; all p< 0.0001). There was no significant difference in the aortic valve area (1.2 ± 0.1 cm^2^ vs. 1.2 ±0.1 cm^2^, p = 0.38). LVEF was significantly higher in the higher AugMAP group (60.6 ± 10.8% vs. 53.3 ± 16.2%, p<0.0001); same as LV end-diastolic volume (135.6 ± 56.8 ml vs. 152.5 ± 67.4 ml, p=0.01) and LV end-systolic volume (65.0 ± 45.8 ml vs. 92.4 ±60.3 ml, p<0.0001). Detailed data are available in **Table 1**

### Survival analysis and Cox regression

During the follow-up period, the all-cause mortality endpoint occurred in 35.2% (n=1525) patients in the higher AugMAP group and 51.5% (n=118) patients in the lower AugMAP group (p<0.0001). Within the first two years, 15.2% (n=692) patients died; and the mortality rate was significantly higher in the lower AugMAP group (13.8% vs. 33.9%, p< 0.0001). In Kaplan-Meier survival analysis, the survival performance differed significantly between the two groups (**Figure 2A**, p<0.0001). In the subsets of patients with normal LVEF and reduced LVEF, we observed a significant difference in the survival curves between the two AugMAP groups (**Figure 2B (p<0.0001) and 2C**, p= 0.0017). In univariate Cox regression, age, presence of CAD, hypertension, diabetes, LV ejection fraction, AugMAP, and Zva were independently associated with all-cause mortality (all p< 0.005). In multivariate Cox regression, the presence of CAD (HR 1.29, 95%CI: 1.16-1.44, p<0.005), DM (HR 1.26, 95%CI: 1.13-1.40, p<0.005) and AugMAP (HR 0.99 per mmHg increase, 95%CI: 0.985-0.993, p<0.005) remained to be independently associated with all-cause mortality. In contrast, Zva became non-significant (HR 1.04, 95%CI: 0.97-1.10, p=0.27) after adjusting for age, sex and LVEF.

**Figure 2.**
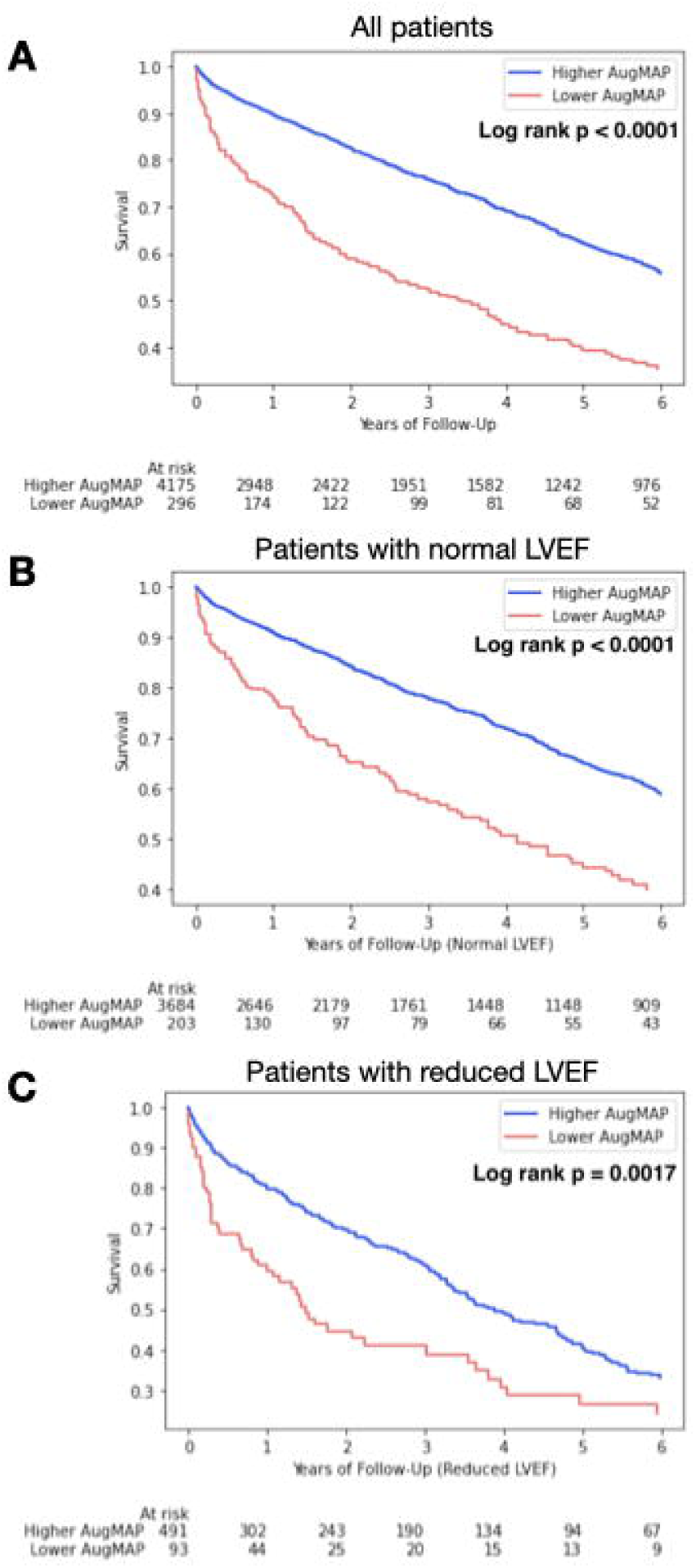
In patients with moderate aortic stenosis, Kaplan-Meier survival analysis showed a significant survival difference between the higher and lower AugMAP groups (cutoff of 80 mmHg). **Panel A**. All patients with moderate aortic stenosis (n=4,563, log-rank p< 0.0001). **Panel B**. Patients with moderate AS and normal LVEF (≥50%), (n=3964, log-rank p<0.0001), and **Panel C**. Patients with moderate AS and reduced LVEF (<50%), (n=599, log-rank p=0.0017).

In all patients with moderate AS, AugMAP was also an independent predictor of all-cause mortality in univariate (HR per unit increase: 0.99, 95%CI: 0.98-0.99, p=0.01) and multivariate Cox regression (HR per unit increase 0.987, 95%CI: 0.978-0.996, p=0.01; adjusted for age, sex and LVEF). Details of univariate and multivariate Cox regression are summarized in **Table 2**.

## Discussion

In this retrospective study, the major findings include 1) augmented MAP can independently predict all-cause mortality in all patients with moderate aortic, regardless of LVEF, 2) patients with low AugMAP had high 1-2 year mortality rate and may benefit more from earlier intervention instead of periodic surveillance. To the best of our knowledge, this is the first study that reported the usefulness of AugMAP in a large moderate AS patient population. Our findings suggest that AugMAP is an important prognostic indicator that can detect subtle LV dysfunction even before the drop of LV ejection fraction. Importantly, AugMAP only requires three input parameters, which minimized the possibility of measurement error and can be easily calculated at the bedside to facilitate the clinical decision-making process. On top of the current aortic stenosis staging system, AugMAP can potentially provide a unified framework for risk stratification in all patients with aortic stenosis.

### The physiological meaning of AugMAP in aortic stenosis

When calculating the augmented blood pressure, we assumed that adding the mean aortic valve gradient to the systemic systolic blood pressure can reflect the actual systolic pressure generated by the left ventricle (12). While noninvasive MAP is practically calculated as the summation of diastolic blood pressure and 1/3 pulse pressure, MAP also equals the product of cardiac output and systemic vascular resistance (SVR) (assuming a negligible central venous pressure level). Therefore, MAP can be considered as the capability of the left ventricle to generate cardiac output in a given systemic vascular resistance, as MAP is proportional to cardiac output when systemic vascular resistance is constant. In the setting of aortic stenosis, a higher augmented MAP indicates better cardiac contractile reserve at rest with better ability to generate blood pressure against the fixed afterload (coupling of the stenotic aortic valve and SVR). The hazard ratio (HR 0.99 per mmHg increase, p=0.01) of AugMAP in Cox regression simply reflects that better contractile reserve should lead to better clinical outcomes. In addition, the shift of optimal cutoff for detecting earlier mortality (**Figure 1A** and **1B**) also implies patients with lower AugMAP are at higher short-to-mid-term mortality risk.

Blood pressure is the most critical part of calculating AugMAP, and potential errors can be as a result of blood pressure control/use of antihypertensive medications and inaccurate blood pressure measurements. In our cohort, the blood pressure in the higher AugMAP group was reasonably controlled. Furthermore, we did not observe a significant difference between the two groups of antihypertensive medications, except beta-blockers and diuretics. While one can question that suboptimal blood pressure control may cause higher blood pressure readings, we would argue that higher blood pressure stands for better cardiac contractile function in patients with moderate AS and can potentially be related to a better outcome. This concept is indirectly supported by an earlier study that patients with higher blood pressure after TAVR procedure are more likely to have LV functional improvement and better clinical outcomes ((14).

### Using augmented MAP to select proper candidates for early intervention: go beyond LVEF

Recently, patients with moderate aortic stenosis and reduced LVEF have gained more interest as this population seems to have an unfavorable prognosis and may benefit more from earlier intervention (3,4,15). Jean et al. have demonstrated that TAVR, instead of SAVR, had significant survival benefits in patients with moderate AS and reduced LV ejection fraction (4). With the ongoing TAVR UNLOAD trial, TAVR indications may further expand to moderate AS patients with reduced LVEF in the near future (5). In the subset of reduced LVEF, we observed an overall similar survival trend of patients compared to published data(3,4,15). Kaplan-Meier survival plot demonstrated a significantly worse survival curve of patients without a subsequent AVR (p<0.0001), supporting the need for earlier intervention instead of periodically surveillance in this population. In our reduced LVEF subgroup, almost 60% of patients with low AugMAP died in a 1-2 year timeframe (**Figure 2C**), which is the current surveillance period suggested by the ACC/AHA guidelines ((1). On top of the present data of moderate AS and reduced LVEF, our data support that AugMAP can further identify patients at even higher risk and may benefit more from earlier intervention.

Beyond the population with moderate AS and reduced LVEF, a large observational study showed that moderate aortic stenosis is associated with increased long-term mortality risk, regardless of LV ejection fraction (15). Thus, while there is no clear evidence for earlier intervention in patients with moderate AS and normal LVEF, it is reasonable to inquire whether these patients would benefit from earlier intervention. Additionally, earlier studies have demonstrated that AVR is not necessarily associated with LV functional improvement. The patients without LVEF improvement had worse clinical outcomes (16,17); these results certainly reflect the need for a prognostic indicator before developing LV dysfunction. In this context, we analyzed the patients with normal LVEF (>50%, n=3,964) in our cohort and found that the same AugMAP cutoff can differentiate the survival performance independent of LVEF (**Figure 2B**, p<0.0001); and there were about 30% accumulative mortality events at two years. This finding suggests that decreased AugMAP can reflect more subtle LV dysfunction even before the LV ejection fraction drops in patients with moderate AS. Our results support the role of AugMAP as an indicator of high-risk moderate AS patients and can potentially be used to select proper candidates of aortic valve replacement independently from LVEF to prevent adverse clinical outcomes.

### A novel risk stratification tool for aortic stenosis

The current ACC/AHA valvular heart disease guidelines suggest using multiple echocardiographic parameters for the staging of aortic stenosis (1). It is commonly encountered that a patient does not entirely fit the diagnosis criteria of a particular stage, so intermediate stages are frequently used to label individual patients in clinical practice properly. Even in the severe stage, LV ejection fraction still needs to be considered to categorize patients with a low-flow, low-gradient condition. Aside from the current staging system, augmented MAP provides a simple and unified framework that integrates cardiac contractile reserve, and the aortic valve mean gradient to assess the overall prognosis independent of LVEF. Requiring only three input parameters minimizes the potential of measurement error and allows this parameter to be easily calculated at the bedside to facilitate the clinical decision-making process.

## Conclusion

AugMAP is a simple and effective novel echo-maker beyond LVEF to identify high-risk moderate aortic stenosis patients who may benefit more from earlier intervention. Our observation demonstrated that AugMAP can detect subtle, early-stage LV dysfunction before the drop of LVEF. On top of the current aortic stenosis staging system, AugMAP can potentially provide a unified framework for risk stratification in all patients with aortic stenosis.

## Limitations

The study is limited by its retrospective, single-center nature. Detailed mortality information was not available to further decipher the exact cause of death. The patient population was predominantly white, so the generalization of our findings needs to be more cautious. The calculation of AugMAP was based on a one-time blood pressure measurement, which can vary time-to-time. Symptom/NYHA class information is not available in our study but will be valuable to further explore the correlations between AugMAP and clinical symptoms. Some patients with reduced LVEF and lower mean aortic valve gradient may have low-flow, low-gradient severe AS. However, we excluded individuals with an aortic valve area of less than 1.0 cm^2^. LV strain data was not available in this cohort, while there might be some correlations between these two parameters gauging LV intrinsic contractility.

Furthermore, stress test data was not routinely available in our study cohort, and effects of aortic stenosis on subendocardial function that were not evaluated. There are no established models to predict the mortality of patients with moderate aortic stenosis, so LV ejection fraction and Zva were used as the reference to assess the parameter performance. The survival performance of SAVR vs. TAVR in this patient population was not tested because only a few patients underwent TAVR in our cohort, reflecting that most of our patient cohort was relatively healthy to tolerate a SAVR procedure.

## Data Availability

NA

## Abbreviations

AS: aortic stenosis
AVG: aortic valve gradient
AugSBP: augmented systolic blood pressure
AugMAP: augmented mean arterial pressure
CO: cardiac output
DBP: diastolic blood pressure
MAP: mean arterial pressure
SBP: systolic blood pressure
SVI: stroke volume index
SAVR: surgical aortic valve replacement
TAVR: transcatheter aortic valve replacement
Zva: valvulo-arterial impedance

## Central illustration

Figure 2. is the representative figure of this manuscript, and is to be served as the central illustration.

## Supplemental Figure

**Suppl. Figure 1.**
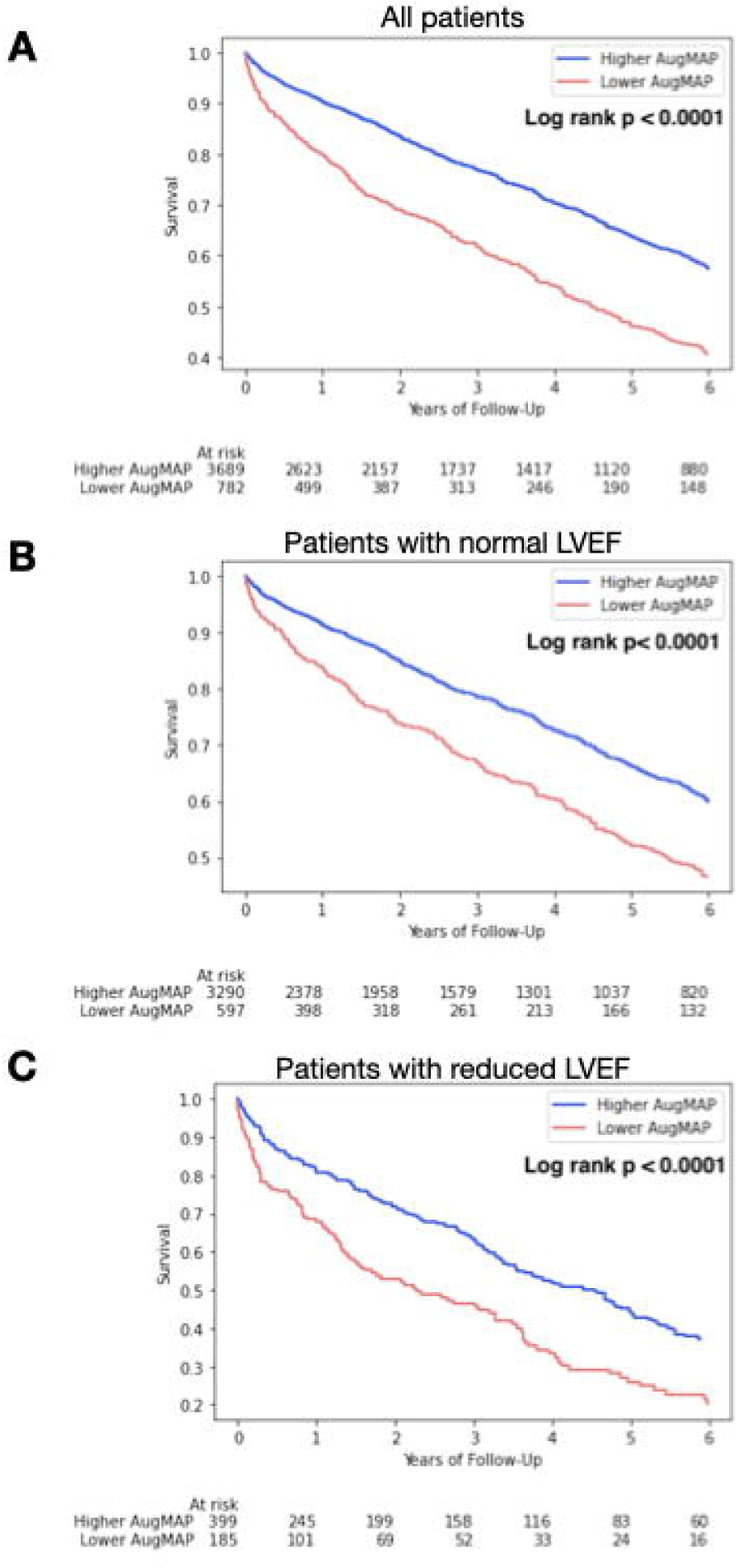
In patients with moderate aortic stenosis, Kaplan-Meier survival analysis showed a significant survival difference between the higher and lower AugMAP groups (cutoff of 87.3 mmHg). **Panel A**. All patients with moderate aortic stenosis (n=4,563, log-rank p< 0.0001). **Panel B**. Patients with moderate AS and normal LVEF (≥50%), (n=3964, log-rank p<0.0001), and **Panel C**. Patients with moderate AS and reduced LVEF (<50%), (n=599, log-rank p<0.0001). Compared to the lower AugMAP cutoff, the separation of survival curves was less apparent in the first two years while the p values reached the same significance.

## Perspectives

Competency in Medical Knowledge: Certain patients with moderate aortic stenosis are at higher risk of mortality and may benefit more from earlier intervention.

Translational Outlook: AugMAP is a simple and effective novel echo-maker beyond LVEF to identify high-risk moderate aortic stenosis patients who may benefit more from earlier intervention.

